# Identifying Heart Failure from Electronic Health Records: A Systematic Evidence Review

**DOI:** 10.1101/2021.02.01.21250933

**Authors:** Rebecca T. Levinson, Jennifer R. Malinowski, Suzette J. Bielinski, Luke V. Rasmussen, Melissa P. Wilson, Sheila M. Manemann, Quinn S. Wells, Veronique L. Roger, Laura K. Wiley

## Abstract

**Background:** Heart failure (HF) is a complex syndrome associated with significant morbidity, mortality, and healthcare costs. Electronic health records (EHRs) are widely used to identify patients with HF. Despite the development of many HF phenotyping algorithms, it is unclear if the characteristics of identified populations reflect the known spectrum of disease in HF patients.

**Methods:** We performed a systematic evidence review to assess the methods used for HF algorithm development from US based data sources and the relevant information facilitating their application to research and clinical care. We queried PubMed for articles published up to November 2021. Out of 428 studies screened, 35 articles were included for primary analysis and 23 studies using only International Classification of Diseases (ICD) codes were evaluated for secondary analysis. Results are reported descriptively.

**Results:** HF algorithms were most often developed at academic medical centers, though these were not evenly distributed across the US. HF and congestive HF were the most frequent labels for observed phenotypes. Diagnoses were the most common data type used to identify HF patients and echocardiography was the second most frequent. Most studies used rule-based algorithms. Validation of algorithms varied considerably with 36% of HF subtype algorithms validated and 67% of acute HF algorithms validated. Demographics of any study population were reported in 57% of algorithm studies and 70% of ICD-only studies, however fewer than half (40% of algorithm and 46% of ICD-only) of studies reported demographics of the HF population identified by their computable phenotype. Of those reporting, most identified majority (>50%) male populations, including both algorithms for HF with preserved ejection fraction identified in this study.

**Conclusion:** There is significant heterogeneity in phenotyping algorithms used to ascertain HF from EHRs. Validation of algorithms is inconsistent and largely relies on manual review of medical records. The concentration of algorithm development at a few sites limits their generalizability to identify HF patients in other settings. Differences between the reported demographics of algorithm-identified HF populations and those expected based on HF epidemiology suggest that current algorithms do not reflect the full spectrum of HF in the general population.

## INTRODUCTION

Electronic health record (EHR) data is increasingly used as a real world data source for clinical and translational research.^1,2^ EHR data are heterogeneous, with differences in both patient populations and data structures making it non-trivial to create robust and reproducible EHR phenotypes. This has limited the scale and portability of phenotyping algorithms ^1,2^. Despite these challenges, EHRs provide an excellent opportunity to identify precise endophenotypes that capture the full spectrum of disease and accelerate the pace of research and clinical translation.

Heart failure (HF) is a complex syndrome that has a heterogeneous presentation,^3^ and is identified through a review of patient history and clinical presentation rather than through a specific test.^4^ The morbidity, mortality,^5,6^ and healthcare costs^7^ associated with HF have triggered significant interest in risk prediction and patient outcome assessment in real world data of clinical populations. The differential response to treatment of multiple subtypes of HF in regards to functional status, left ventricular ejection fraction (EF) level, or clinical stage ^3,8^; as well as subphenotypes identified through clustering of clinical trial data, support the need for enhanced phenotyping of HF populations ^9,10^. As such, EHRs have long been considered a prime resource for research into the phenomics HF.

However, despite more than a decade of work, how to ascertain HF from EHRs remains an area of active research.^11,12^ The complexity of both the HF phenotype and the EHRs in which computational approaches at defining that were implemented has led to numerous publications of HF EHR phenotyping algorithms without the emergence of a single consensus algorithm, even as the idea of a universal definition of HF in the clinical setting has gained traction.^13^ Several studies have developed or compared algorithms to computationally identify HF populations in the EHR.^14,15^ However, it remains unclear whether populations identified by existing EHR algorithms represent the full spectrum of HF in the general population. Furthermore, when evaluating individual algorithms it can be difficult to understand what type(s) of HF patients are being captured, thereby limiting the ability to determine if the results can be generalized to other populations, essential information for evaluating any evidence generated from these patient groups.

To elucidate these issues and provide a rational foundation for future phenomics projects,^16,17^ we performed a systematic evidence review to evaluate how HF phenotypes are identified from the EHR and reported in the literature. We discuss active difficulties in the phenomics literature of HF. We compare these EHR algorithms and their component parts. We also assess their utility for research and clinical application, by comparing characteristics of algorithm defined populations to the general population. We show that for HF phenomics to benefit all HF patients, we need to carefully consider both the context, context, and results of EHR phenotype algorithms.

## METHODS

### Search Strategy

Search terms were refined with an iterative process involving co-authors RTL, SJB, LVR, JRM, and LKW and incorporated Medical Subject Heading (MeSH) terminology to identify relevant secondary subject headings. This systematic evidence review (SER) was conducted as a subanalysis of a larger review investigating the quality of EHR-based cohort identification algorithms and was not registered.Accordingly all studies were first captured by the search string used by the larger SER:

> ((((electronic health records OR “EHR” OR electronic medical records OR “EMR”) AND (“natural language processing” OR “machine learning” OR classifier OR “deep learning” OR “artificial intelligence” OR phenotyp* OR “phenome” OR ICD OR probabilistic OR algorithm OR (“data-mining” OR “data mining”)))))

Studies were further refined to only include those relevant to heart failure by applying the following search string:

> (((((((“heart failure”[MeSH Major Topic]) OR ((cardiac or myocardial) AND (failure or insufficiency))) OR (congestive heart failure)) OR (congestive cardiomyopathy)) OR (cardiomyopath*)) OR ((cardi* or heart* or myocard*) AND (fail* or incompet* or insufficien* or decomp*)))

Search results for the larger project were uploaded to Covidence, a software for SER management. Heart failure search results were identified and prioritized for screening, review, and data extraction for this study. Importantly, if a study labeled the phenotype identified as heart failure, we kept this label, regardless of the clinical criteria within the computed phenotype.

### Article Review

Search results were assessed in Covidence for inclusion in duplicate by blinded reviewers according to a pre-specified protocol. Conflicts were adjudicated through discussion or the input of a third reviewer, as necessary. Studies were included if they used EHR data to derive a phenotype that was a disease or condition, disease subtype, or disease symptom. Non-primary research articles, non-human studies, or those that were performed outside of the United States or those that only used claims data were excluded. Studies that did not describe identifying a patient population, identified the final patient population through International Classification of Disease Clinical Modification (ICD) diagnosis codes only, manual chart review, or simply applied previously published algorithms were also excluded. We reviewed all studies excluded for using a previously published algorithm to extract the algorithm citation and add to the overall study list if not already captured in our search strategy.

For this subanalysis we excluded those studies that did not identify a phenotype labeled as HF or a HF subtype, and we performed a secondary extraction of those studies excluded from the larger study that described extracting the HF population using only ICD diagnosis codes.

### Data Extraction

Reviewers (RTL, SJB, LVR, JRM, MPW, SMM, LKW) extracted data in duplicate from included articles blinded to the other’s responses using a two-stage process. First we extracted information about the focus of the article (i.e., algorithm method development, applied study, or portability of previously published algorithm method) defining applied studies to be those where the primary purpose of the paper was to investigate a scientific question or describe a clinical application, rather than developing an algorithm to identify HF patients. We also extracted the label used by the author for all eligible phenotypes (e.g., “congestive heart failure”, “acute congestive heart failure”, and “heart failure” were all considered to be different phenotypes). Discordance between reviewers’ responses were resolved through discussion or by author LKW or JRM as a third reviewer. Next, we extracted additional information about each study and algorithm into custom forms. Data extraction focused on identifying study details (e.g., study data source, type of applied study, etc.), algorithm methodology (e.g., type of data, type of computational method, etc.), algorithm validation (e.g., type of validation, criteria for determining HF during manual review, etc.), and algorithm generalizability (e.g., demographics reporting, etc.). For papers that reported development of algorithm/s for multiple phenotypes we extracted algorithm specific information independently for each qualifying phenotype. Neither quality assessment nor risk of bias were assessed for the included studies, as there is no validated instrument for these types of studies.

We conducted additional data extraction for information specific to HF. For algorithm-based studies, we extracted who performed chart review validation and demographic information (sex, age, race/ethnicity) for the population identified by the algorithm where available. To understand the relationship between algorithm-identified HF populations and those identified using only ICD diagnosis codes, we also performed a minimal data extraction on studies excluded in the full text review as ICD code only. This limited analysis extracted the type of study, ICD-only phenotypes, whether the ICD definition was validated, and demographic information (sex, age, race/ethnicity) for the population identified by the algorithm where available. These HF specific data extractions were performed via co-extraction by RTL and LKW.

### Analysis Plan

We analyzed and reported data extracted using descriptive statistics (counts, etc.) and visualization. We manually classified journals as “informatics” (e.g., focused on biomedical informatics or computer science topics) or “clinical” (e.g., focused on clinical domains or healthcare delivery). We harmonized the EHR data source reported by each study (e.g., “Mayo Clinic EHR” and “Mayo Clinic hospital” harmonized to “Mayo Clinic”), classified the data source type (i.e., “Academic Medical Center”, “Data Clearinghouse”, “Health Information Exchange”, “Regional Healthcare System”, “Veterans Affairs”, or “Other”), and identified the primary state where the EHR data source was located. For those studies reporting the definition used during manual review of patient records, two cardiologists (VLR, QSW) classified the provided definition by whether it met Framingham^18^, ARIC^19^, or another standard diagnostic criteria. We performed all analyses using R version 3.6.0^20^ and a variety of packages for data processing, graphing, and reporting.^21–29^

## RESULTS

The initial literature search for the primary review was performed on January 29, 2019 and updated on November 11, 2021. Of the 5,946 studies assessed in the primary review, 418 were selected for this subanalysis using the query to identify HF specific studies. A further 14 studies that were identified from algorithm citations were added to the screening list. After removing duplicates, a total of 428 studies were screened for inclusion with 227 (53.0%) continuing to full text review. Full text review excluded 169 (74%) of the studies (Supplemental Table 1), with the majority excluded for not identifying HF (N=44), identifying the phenotype using chart review (N = 31), or not identifying a patient population (n=27). **Figure 1** shows the PRISMA flow diagram providing a full accounting of all studies reviewed, excluded, and included in the final subanalysis.

**Figure 1.**
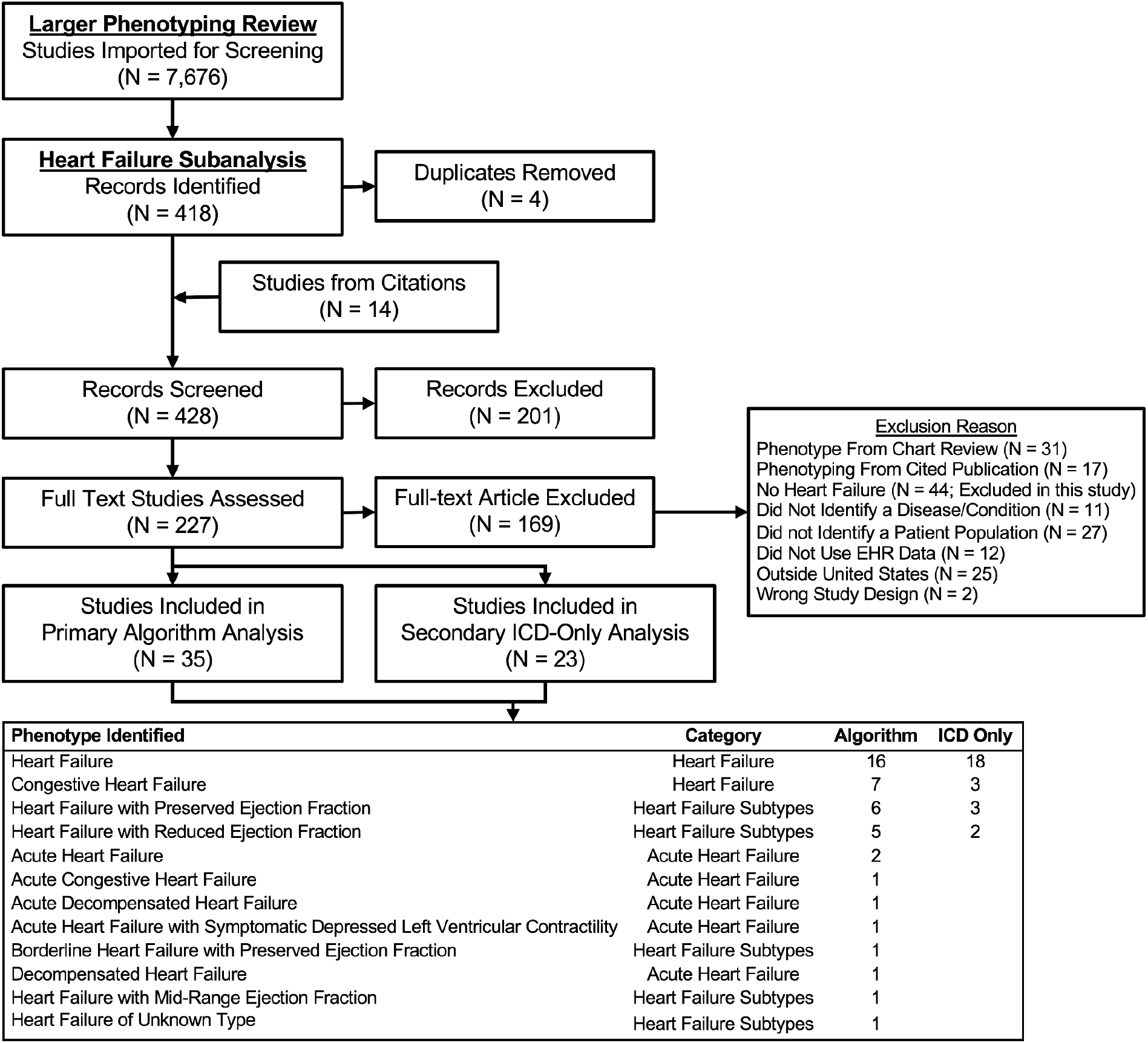
PRISMA Diagram.

58 studies met criteria for data extraction (Supplemental Tables 2-4). These studies contained a total of 69 computed phenotypes, 43 identified with algorithms and 26 defined using only ICD codes. Across all study types, “Heart failure” (N = 34), “Heart Failure with Preserved Ejection Fraction” (N = 9), “Heart Failure with Reduced Ejection Fraction” (N = 7), and “Congestive Heart Failure” (N = 10) were the most frequent phenotypic labels identified. We further categorized these phenotypes into all-cause “Heart Failure”, “Acute Heart Failure”, “Heart Failure Subtypes”. **Figure 1** provides a complete list and frequency of phenotypes identified by study type. We further categorized these phenotypes into all-cause “Heart Failure”, “Acute Heart Failure”, “Heart Failure Subtypes”

### Heart Failure Detection Methods

To better understand how studies identified HF populations from EHRs we assessed the types of data and methods applied across the 43 algorithm identified phenotypes.The majority of phenotype algorithms (91%, N = 39) used two or more different types of data to identify HF patients. Diagnoses were the most common data type used across algorithms (93%, N = 40). Of those algorithms using diagnoses, 73% used them alone (N = 29) or in combination with SNOMED or other terminologies (20%, N = 8). Echocardiography data - either the presence of a report or extracted ejection fraction values - were the second most frequent data type (58%, N = 25) and were used in 39% of algorithms detecting HF (N = 9), 93% of algorithms detecting HF subtypes (N = 13), and 50% of algorithms detecting acute HF (N = 3). **Figure 2A** reports the frequency of each data type and data type combinations used across study phenotypes.

**Figure 2.**
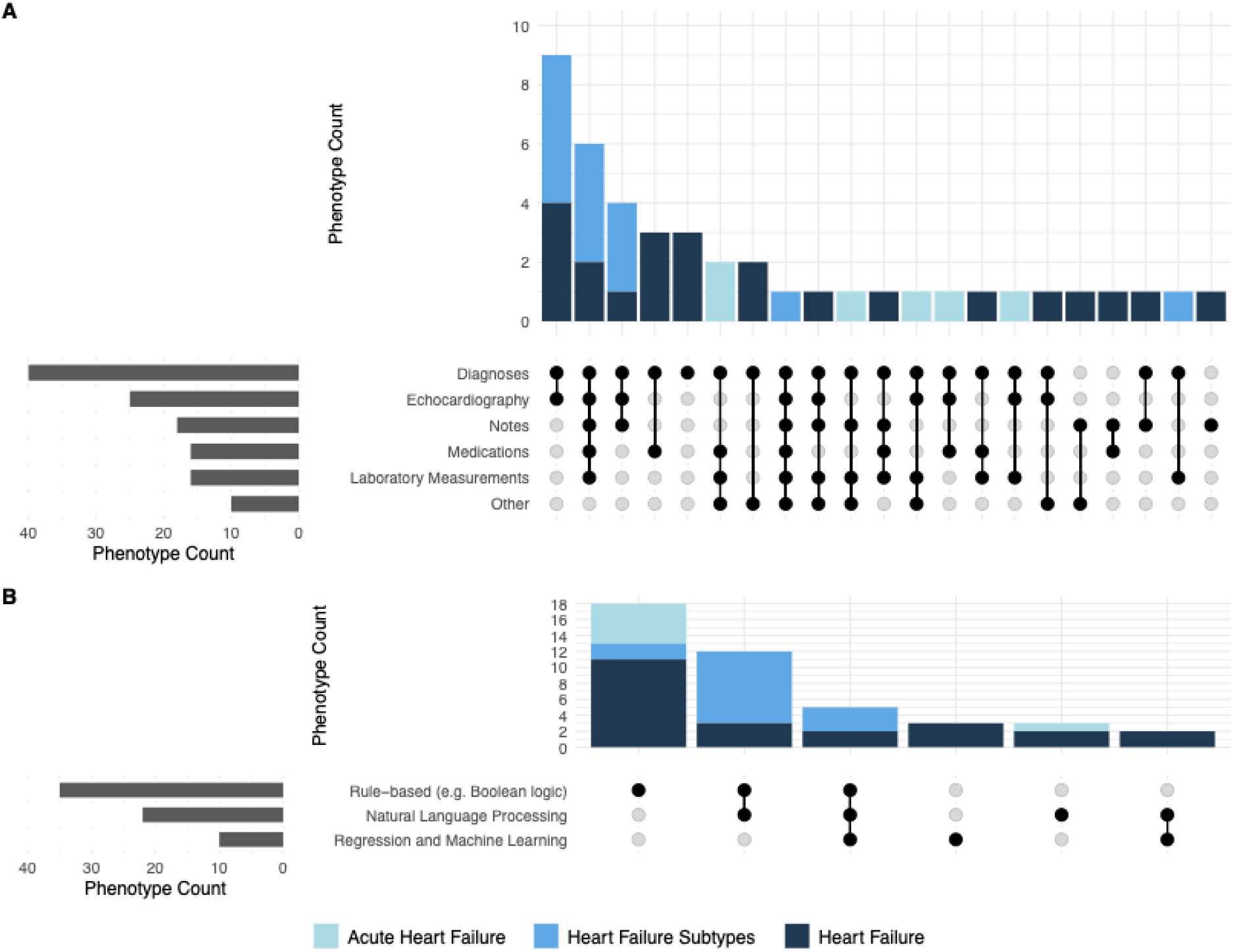
Algorithm Data Sources and Methods. (A) Types of electronic health record (EHR) data used across the evaluated algorithm identified phenotypes (n=43). Black dots connected by lines show where multiple data sources are used (e.g., the first bar shows that both Diagnoses and Echocardiography were used); (B) Frequency of algorithm methods used across the evaluated phenotypes (n=43). Black dots connected by lines show where multiple methods were used (e.g., the second bar shows algorithms that used both Rule-based and Natural Language Processing methods).

The majority of study phenotypes used some combination of rule-based methods (81%, N = 35), of which 49% (N=17) also used text extraction or natural language processing. A minority of study phenotypes were identified using regression or machine learning methods (23%, N = 10). None of the studies with acute HF as a phenotype used regression or machine learning methods. **Figure 2B** reports the frequency of each methodology and methodology combinations used across study phenotypes.

### Heart Failure Definition and Validation

To understand the potential applicability of each identified study phenotype for cardiovascular research, we assessed whether and how the ICD definition or algorithm were validated. **Figure 3A** reports the frequency of validation across all study phenotypes. Overall, only a third of the algorithms were validated. Of these, 52 % (N = 12) of HF algorithms, 36% (N = 5) of HF subtype algorithms, and 67% (N = 4) of acute HF algorithms were validated. The majority of validated algorithms used manual abstraction of patient records (81%, N = 17) with 76% (N = 13) of them providing additional detail on how HF was defined during abstraction. **Figure 3B** reports the frequency of established criteria^18,19^ used to define HF during abstraction. Of the 13 validated algorithms that provided additional information about their criteria in abstraction, 69% (N=9) used the Framingham HF Diagnostic criteria, either alone or in combination with other information, to determine HF status.

**Figure 3.**
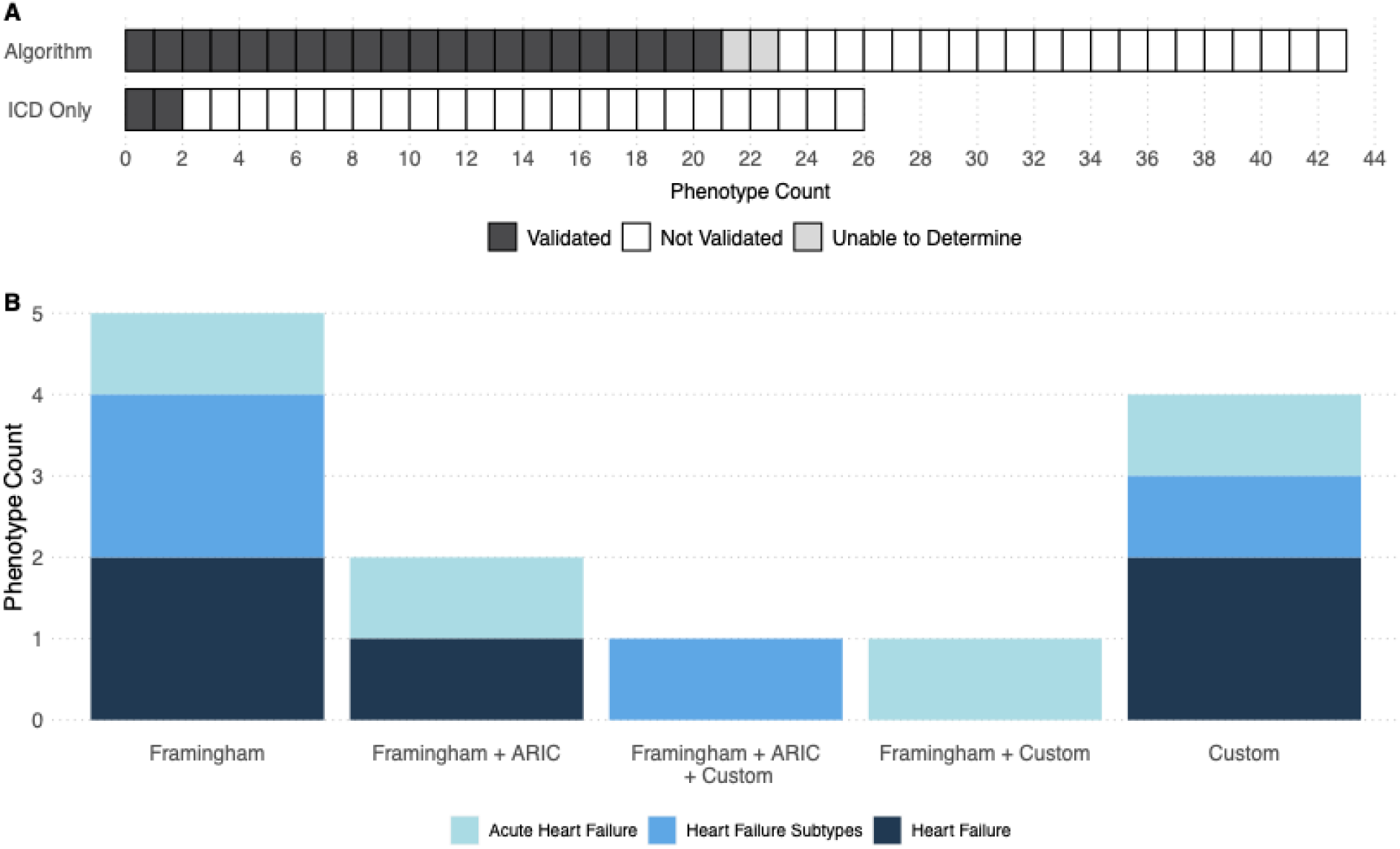
Heart Failure Definition and Validation. (A)Number of phenotype algorithms that were validated. This includes the algorithm-based studies in our primary analysis (n=43), as well as the sub-analysis of studies that only used International Classification of Diseases (ICD) codes (n=26); (B) Frequency of review criteria used in the validation of phenotype algorithms, for the 13 phenotype algorithms that described their validation criteria. Studies may use a single criterion, or may have used a combination of criteria. Framingham = Framingham Heart Failure Diagnostic Criteria, ARIC = Atherosclerosis Risk in Communities, Custom = study-specific definition.

### Algorithm Development and Application

To better understand how HF algorithms and ICD-only definitions have been presented in the literature, we analyzed study types and publication trends. **Figure 4A** reports the frequency of study type published over time separated by whether the study used an algorithm or an ICD-only definition. The computed phenotypes identified in this analysis were fairly evenly split between algorithm method development (N = 30) and applied (N = 39). However more than half (61%, N = 26) of algorithm studies were method development focused, while more than three quarters (85%, N = 22) of ICD only studies were applied.

**Figure 4.**
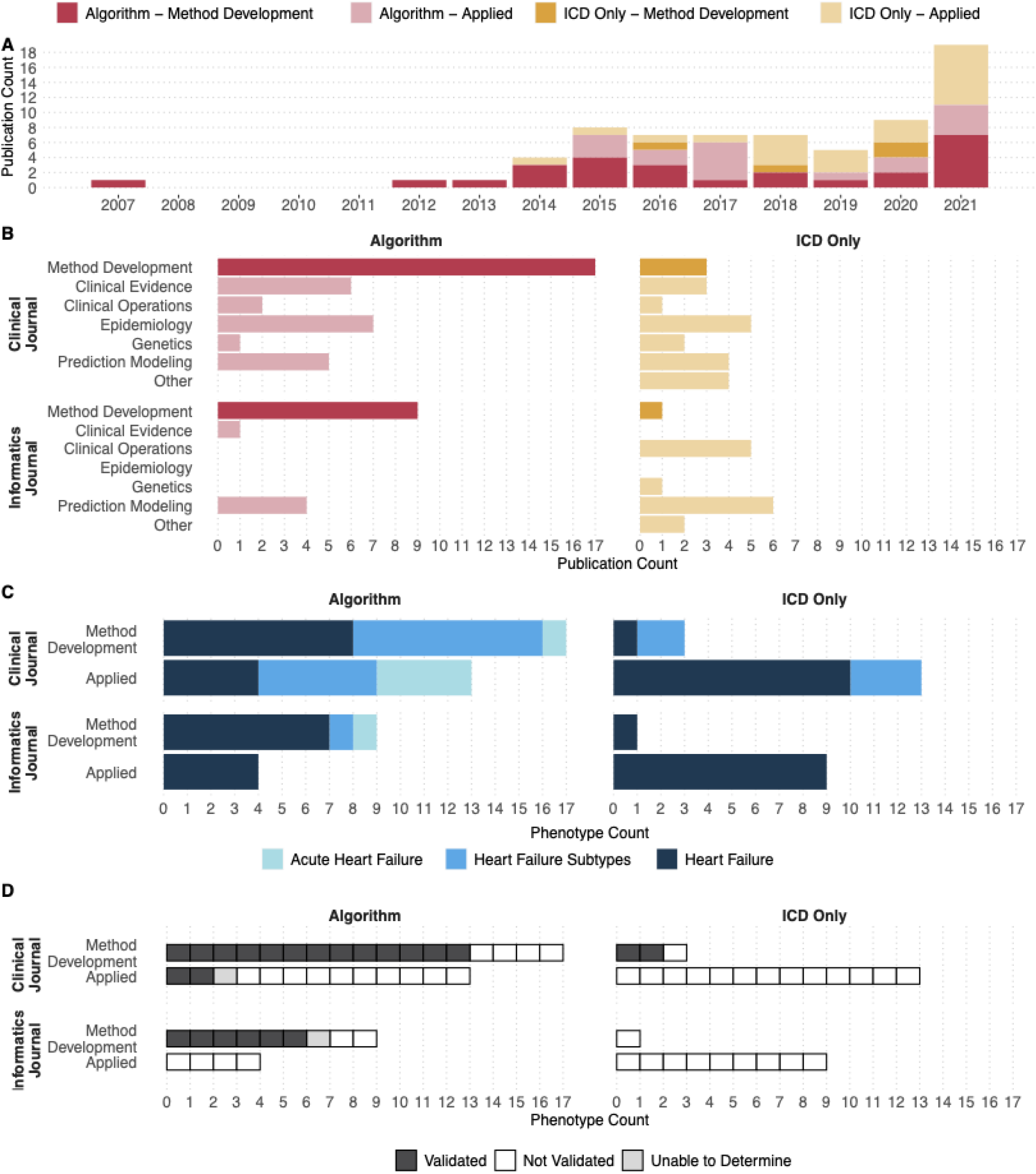
Algorithm Development and Application. (A) Year of publication of analyzed studies, which includes the primary analysis set of phenotype algorithms (n=43) and the secondary analysis set of studies that used only International Classification of Diseases (ICD) codes (n=26). Studies are further subdivided if the primary focus was on algorithm method development or its application; (B) Frequency of studies published in a primarily clinical journal vs. a primarily informatics journal based on the study type (algorithm method development or type of applied study) for both algorithm identified phenotypes and ICD-only phenotypes. ; (C) Frequency of the study focus (method development vs. applied) across journal type (clinical vs. informatics), stratified by heart failure class for the primary phenotypes analyzed (n=30); D) Frequency of validation pf phenotype definition by journal type (clinical vs. informatics) and study focus (method development vs. applied).

Next we analyzed the types of journals publishing these studies. The majority of studies identified in this analysis were published in clinical journals (67%, N = 46). **Figure 4B** reports the frequency of algorithm and study types presented in each type of journal. We further classified each applied study by all of the application domains (i.e., clinical evidence, clinical operations, epidemiology, prediction modeling, other, plus any combination). The most common applied study type was prediction modeling (N = 19). The most frequent combination was prediction modeling applied to clinical operations (e.g., creating dashboard, clinical decision support, etc.) (N = 6), with all of these studies using ICD only definitions of HF and the majority publishing in informatics journals (N = 5).

Among computed phenotypes for HF we also considered whether algorithm, ICD only, or validation status differed by study or publication type. About half of HF algorithms were published in clinical journals (52%, N = 12), while a larger majority of acute HF (83%, N = 5) and HF subtype (93%, N = 13) algorithms were published in clinical journals. Within informatics journals the two algorithms identifying acute HF and HF subtypes were both part of method development papers. Within clinical journals the type of study (e.g., method development vs. applied) was predominantly applied within acute HF (N = 4 Applied, N = 1 Method Development) and predominantly methods development within HF subtype (N = 5 Applied, N = 8 Method Development) algorithms. All HF subtype algorithms used in applied studies (N = 5) were used for a combination of clinical evidence, prediction modeling and epidemiology, while all HF algorithms used in applied studies published in informatics journals (N = 4) were used for prediction modeling. **Figure 4C** reports the frequency of phenotype algorithm types by study, journal type, and phenotype label. Among algorithms defined in method development papers, 77% (N = 13) of those published in clinical journals were validated, compared to only 67% (N = 6) of those published in informatics journals. Validation in applied studies was rare, with only 2 instances of validation across 17 published algorithms. **Figure 4D** reports the frequency of algorithm validation by study and journal type.

### Algorithm Generalizability

To better understand the potential generalizability of the HF algorithms identified we analyzed the locations and types of clinical data sources used to create each algorithm. The most common phenotypic data sources were academic medical centers (N = 19). **Figure 5A** reports the frequency of data source type for algorithm derived phenotypes. Although 11 individual states were represented in the literature, the majority contributed to only a single algorithm. Minnesota was the most common site of algorithm development publishing 11 algorithms, with the majority coming from the Mayo Clinic (N = 8). A total of 15 algorithms were developed with data from multiple states, the majority from the VA (N = 5) and Kaiser Permanente (N = 5). Additionally, 6 algorithms either did not list the EHR source at all (N = 5) or did not provide sufficient details about the EHR source to identify a location (N = 1). **Figure 5B** reports the frequency of data source location for algorithm derived phenotypes.

**Figure 5.**
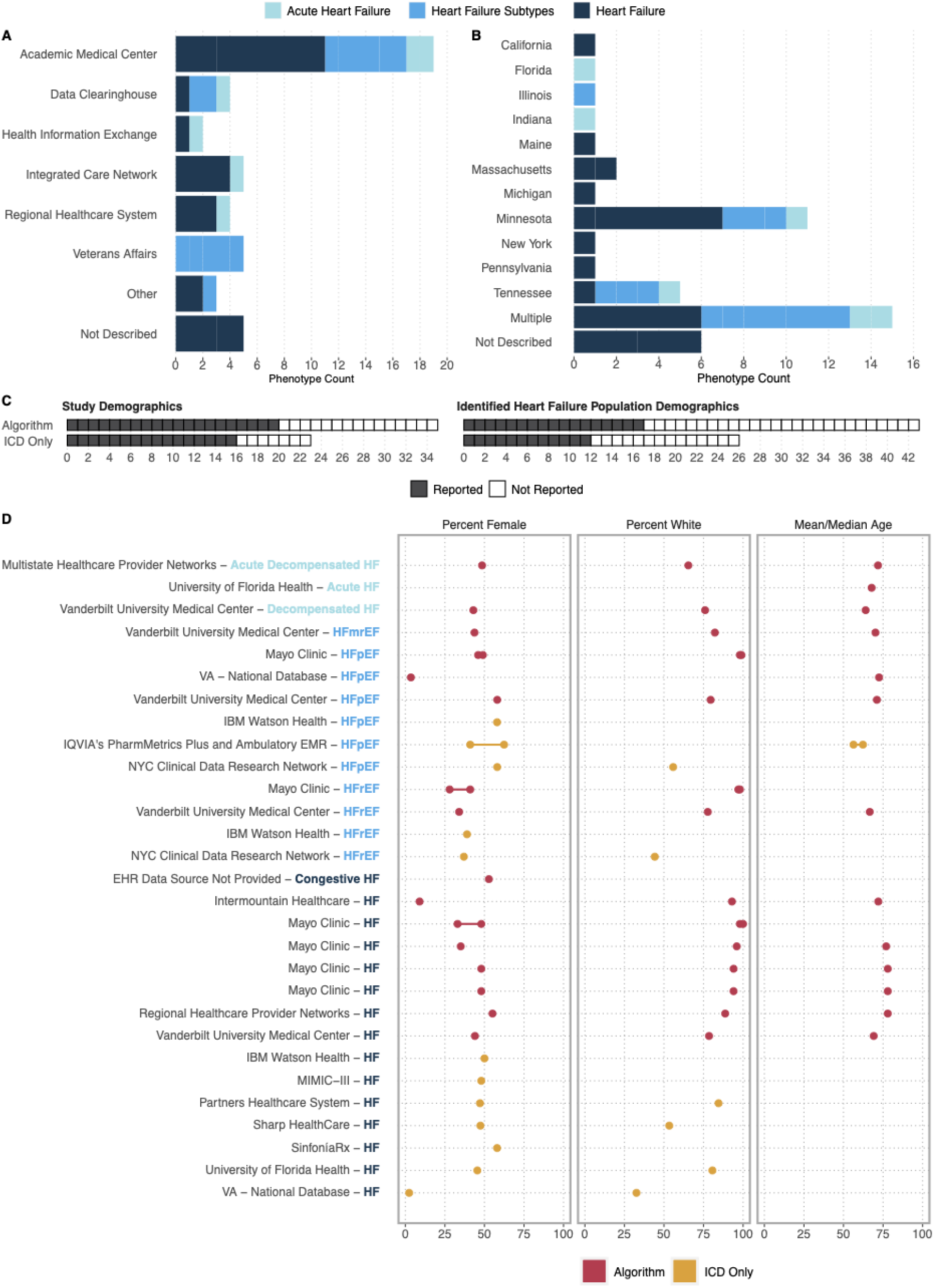
Study Population Characteristics. (A) Frequency of EHR data source and (B) geographic location of the EHR data source population used to create each phenotype algorithm included in the primary analysis (n=43); (C) Frequency of the primary analysis algorithm definitions (n=43) and subanalysis International Classification of Diseases (ICD)-only phenotypes (n=26) that report any study demographics or demographics of the phenotype-identified heart failure population; (D) Phenotype description, data source, and reported demographics for both algorithm and ICD-defined phenotype definitions (when reported).

Understanding the population captured by an algorithm is essential to evaluating its generalizability. Therefore, we investigated which demographics were reported by each study. Demographics of any study population (not necessarily the identified HF population) were provided in only 57% (N = 20) of algorithm studies and 70% (N = 16) of ICD only studies. Regardless of HF identification approach, reporting study demographics was more common among studies published in clinical journals (Algorithm: 59%, N = 13; ICD Only: 85%, N = 11) compared to informatics journals (Algorithm: 54%, N = 7; ICD Only: 50%, N = 5).

The minority of studies provided demographics for the identified HF populations, though reporting was less common among algorithm-defined phenotypes (40%, N = 17) compared to ICD only phenotypes (46%, N = 12). **Figure 5C** reports the frequency of reporting study and HF population demographics across algorithm and ICD only studies. For those HF populations with demographic reporting, 97% reported patient sex (N = 28), 69.0% reported frequency of at least one race or ethnicity (N = 20), and 48% reported mean or median age of the population (N = 14). The majority (N = 20; 71% of studies reporting sex) of HF definitions identified majority male populations (>50% male) including both algorithms detecting HF with preserved ejection fraction. **Figure 5D** reports the phenotype, data source, and reported demographics for available algorithm and ICD identified HF populations.

## DISCUSSION

Our results demonstrate that while EHR-based phenotyping of HF has been an active area of research for many years, it remains an open area with multiple unexplored domains. HF phenomics dates back more than a decade, and has produced at least one published method development focused algorithm every year since 2012. There appears to be an opportunity to identify more complex phenotypes as the majority of identified algorithms detected all-cause HF (e.g., with a minority identifying specific HF subtypes or acute HF events. However even within all-cause HF, there remains significant heterogeneity in the types of data used across algorithms, suggesting that even broadscale patient identification is not yet a settled task. Finally, although prediction modeling was a common application domain for HF populations, few of the identified studies actually used machine learning within their phenotyping algorithms. It is unclear whether this finding is due to limitations of our search strategy, a reflection of positive publication bias (e.g., machine learning methods have been applied, but performed poorly and were not published), or a true gap in the literature.

Nevertheless, investigation of machine learning algorithms may present a rich opportunity for HF phenomics research.

Another area of potential investigation is in the utility of HF algorithm performance for research and clinical applications, though this effort was hampered by the moderate rates of algorithm validation. Although one can use the logical combination of data types in algorithms to infer the patient population identified, validation serves an important role in tuning algorithm performance to the types of levels of evidence used during chart review to create a gold-standard case label. Just as clinical trial inclusion/exclusion criteria affect the generalizability of findings to the clinic, the phenotype definition used during validation determines whether an algorithm is fit for a particular analytic purpose. Positively, the majority of studies performing validation included the phenotype definition used during chart review. Interestingly, two used clinical trial definitions, which suggests the potential utility of these algorithms to demonstrate generalizability of trial findings in EHR populations.

Investigators seeking to apply published algorithms should pay close attention to the phenotype definition used during validation to determine if the identified population is appropriate for the intended analysis.

We also considered whether the populations identified by published HF algorithms capture the full spectrum of HF patients. We see significant potential for selection bias given the reliance on echocardiography by the majority of algorithms. For example, some studies labeled the phenotype they were capturing as “Heart failure”, but required an EF <40 in the algorithm meaning that the algorithm could actually only identify reduced EF heart failure. Furthermore, the majority of algorithms were developed at tertiary care academic medical centers where providers often rely on outside reports of ejection fraction measurements. These outside reports are rarely integrated into clinical data warehouses potentially affecting performance of algorithms across institutions. Therefore, it is possible that a number of HF patients are not being captured by these algorithms. Similarly, the concentration of algorithm development within a single state (Minnesota) and even a single institution (Mayo Clinic) or in institutions with non-representative populations (e.g., VA) limits the generalizability of these algorithms to community clinics and populations in underrepresented regions. As expected, algorithms developed at institutions in low-diversity geographic regions identify majority white populations, while the VA identified nearly 100% male populations. However even after accounting for these expected deviations, there remains stark differences from demographics expectations, with only two algorithm and one ICD studies identifying majority female populations despite the prevalence of HF in the United States being majority female (52%).^30^ Large scale projects like the National Heart Lung and Blood Institute’s HeartShare program should be particularly mindful of these concerns to ensure all HF patients benefit from their phenomics investment.

Our study also shines light on potential challenges in dissemination and implementation of HF phenomics. First, we were surprised to find the majority of complex phenotypes published in clinical journals. Existing reviews of phenotyping methodology have focused solely on informatics and computer science journals,^31,32^ an approach which our findings suggest may miss a number of complex algorithms. Similarly we expected more applied studies to be published in clinical journals, whereas our findings show the majority were published in informatics journals. We also noticed important trends in terms of the use of algorithms vs. ICD only definitions across application domains. In areas where codes are more commonly used (e.g., claims data), or where HF was not the primary interest area (e.g., computer science development with an HF application) we saw more applied studies using ICD only HF definitions. However, we see a potential concern for implementation of clinical evidence created using HF phenomics: although most clinical evidence studies used algorithms, the majority of clinical operations studies (e.g., clinical decision support or dashboard operations) used ICD only definitions. It will be critical to ensure alignment between the patient populations used to develop treatment guidelines and those for whom EHRs can implement practice guidance.

This study also identified significant areas of opportunity for the field to improve the rigor and reproducibility of both algorithm development and the reporting of algorithm-based studies. Our investigative team with decades of combined experience in EHR-based phenotyping were unable to fully identify the data sources, methods used, or validation status for five algorithms. A third of algorithm-based studies and nearly half of ICD only studies reported no demographics for any population in the paper. While publishing the demographics of algorithm identified populations has not historically been expected, the lack of reporting limits the ability to determine the potential generalizability of the algorithms. Especially with a phenotype as complex as HF, demographics provide important insight into the specific subpopulations that an algorithm may be preferentially identifying.

A major strength of this study is the unbiased manner by which we identified papers containing HF algorithms. However it is also possible that our search strategy did not identify all studies even if they contained algorithms that would have met criteria for inclusion. The lack of standard definitions and terminology for cohort identification^31^ make complete ascertainment of phenotyping studies difficult.

However, we used an interactive process with feedback and positive controls from the full author group to ensure relevant papers were included. Additionally, while assessing the risk of bias or study quality is an essential component of SERs, no tool currently exists to measure the risk of bias for EHR phenotyping algorithm studies. In absence of such a framework, we are unable to assess the risk of bias or quality of the included literature beyond noting the poor reporting of basic study information (e.g., where the population is derived).

In conclusion, we have completed the first systematic evidence review of computational phenotyping in HF. Overall our data suggest that current EHR identified HF populations do not reflect the full spectrum of HF phenotypes or patient populations. We also found significant room for improvement in reporting of HF phenotyping algorithms and study populations. There appears to be significant opportunities to advance the field of HF phenomics in both phenotypic and algorithm complexity moving forward.

## Data Availability

Data will be made available at the time of publication.

## Acknowledgements

We would like to thank David Mayer for providing project management support

